# Towards cascading genetic risk in Alzheimer’s disease

**DOI:** 10.1101/2023.12.16.23300062

**Authors:** Andre Altmann, Leon M Aksman, Neil P Oxtoby, Alexandra Young, ADNI, Daniel C Alexander, Frederik Barkhof, Maryam Shoai, John Hardy, Jonathan M Schott

## Abstract

Alzheimer’s disease typically progresses in stages, which have been defined by the presence of disease-specific biomarkers: Amyloid (A), Tau (T) and neurodegeneration (N). This progression of biomarkers has been condensed into the ATN framework, where each of the biomarkers can be either positive (+) or negative (-). Over the past decades genome wide association studies have implicated about 90 different loci involved with the development of late onset Alzheimer’s disease. Here we investigate whether genetic risk for Alzheimer’s disease contributes equally to the progression in different disease stages or whether it exhibits a stage-dependent effect.

Amyloid (A) and tau (T) status was defined using a combination of available PET and CSF biomarkers in the Alzheimer’s Disease Neuroimaging Initiative cohort. In 312 participants with biomarker-confirmed A-T- status, we employed Cox proportional hazards models to estimate the contribution of *APOE* and polygenic risk scores (beyond *APOE*) to convert to A+T- status (65 conversions). Furthermore, we repeated the analysis in 290 participants with A+T- status and investigated the genetic contribution to conversion to A+T+ (45 conversions). Both survival analyses were adjusted for age, sex, and years of education.

For progression from A-T- to A+T-, APOE-e4 burden showed significant effect (HR=2.88; 95% CI: 1.70-4.89; P<0.001), while polygenic risk did not (HR=1.09; 95% CI: 0.84-1.42; P=0.53). Conversely, for the transition from A+T- to A+T+, the APOE-e4 burden contribution was reduced (HR=1.62 95% CI: 1.05-2.51; P=0.031), while the polygenic risk showed an increased contribution (HR=1.73; 95% CI:1.27-2.36; P<0.001). The marginal *APOE* effect was driven by e4 homozygotes (HR=2.58; 95% CI: 1.05-6.35; P=0.039) as opposed to e4 heterozygotes (HR=1.74; 95% CI: 0.87-3.49; P=0.12).

The genetic risk for late-onset Alzheimer’s disease unfolds in a disease stage-dependent fashion. A better understanding of the interplay between disease stage and genetic risk can lead to a more mechanistic understanding of transition between ATN stages, a better understanding of the molecular processes leading to Alzheimer’s disease as well as opening therapeutic windows for targeted interventions.

## Introduction

Alzheimer’s disease is characterized, on the neuropathological level, by the buildup of two proteins: amyloid plaques and neurofibrillary tangles of phosphorylated tau^1^. Both these pathological features can be observed long before the memory loss and decline in executive function that is characteristic of patients with Alzheimer’s disease. Accumulation of the amyloid plaques in the brain predates the clinical symptoms of Alzheimer’s disease by two decades^2^, while the spatial distribution of tau tangles reflects more closely the reported cognitive deficits and neurodegeneration^3^.

The amyloid cascade hypothesis postulates that the deposition of the amyloid-β protein – the main component of the amyloid plaques – is the cause of Alzheimer’s disease, and that neurofibrillary tangles, cell loss, vascular damage and dementia are a direct consequence^4^. In keeping with the amyloid cascade hypothesis, a theoretical framework for the progression of biomarkers during the course of Alzheimer’s disease has been developed^5^. Here, amyloid pathology is the first to appear, followed by tau pathology, neurodegeneration and finally cognitive decline. Support for this theoretical framework comes from a number of lines of evidence, including a variety of data-driven modeling approaches based on biomarker data ^6–10^. In an attempt to operationalize this, a simplified ATN model has been proposed^11^. The components of the ATN model refer to the status of three different key biomarkers in Alzheimer’s disease: amyloid (A), tau (T) and neurodegeneration (N). In this approach, each of the three biomarkers can either be positive or negative. That is, exceeding the centiloid threshold in amyloid PET imaging would place a participant into the A positive (A+) group, while a scan just below the threshold would be considered amyloid negative (A-). One practical advantage (but also a major source of criticism) of this model is that the biomarker status can be assessed using a variety of methods: wet biomarkers (CSF or plasma) or brain imaging (centiloids or visual reads)^12^. Although the progression from A-T-N- to A+T-N- to A+T+N- to A+T+N+ would be the most typical progression in Alzheimer’s disease and in keeping with the theoretical framework of biomarker progression, all combinations of biomarker statuses emerge in observational cohorts^13,14^.

Genome-wide association studies (GWAS) have broadened the understanding of the genetic basis of late-onset Alzheimer’s disease (LOAD) over the last decades^15^. Initially, these studies were restricted to cases with a clinical diagnosis of Alzheimer’s disease and healthy controls^16–18^. Recently, these GWAS have been enriched with participants with a family history of Alzheimer’s disease (diagnosis-by-proxy)^19–21^, leading to a drastic increase in sample size and expanding the set of genetic risk loci for Alzheimer’s disease to 90^15^. These loci have been linked to various molecular processes such as immunity, cholesterol processing and endocytosis^22^. Further studies investigated the genetic effects on Alzheimer’s disease-related biomarkers ranging from tau and amyloid levels in CSF^23,24^, in the brain^25,26^ to MRI-based measures such as hippocampal volume^27^ and phenotypes derived from disease progression modeling^28^. The strongest common genetic risk factor for Alzheimer’s disease is the e4 allele of the *APOE* gene: carriers of the e4 allele have a 2-4-fold increased risk to develop Alzheimer’s disease, while e4 homozygous subjects have an 8-12-fold increased risk^29^. The genetic risk outside the *APOE* region is often summarized using polygenic scores, which have been shown to improve predictions of clinical diagnosis^30^ and pathology-confirmed cases^31^. Likewise, the effect of *APOE* as well as the polygenic risk on various imaging and non-imaging biomarkers have been investigated^32–35^, with ongoing work suggesting that risk accumulated along different molecular pathways exerts differential effects on different biomarkers in Alzheimer’s disease^35–37^.

For Alzheimer’s disease and for other disorders, genetic risk is often considered as a time-invariant constant. That is, genetic risk identified through case-controls studies is assumed to affect both onset and progression. However, as Alzheimer’s disease is now understood to unfold in stages, we hypothesized that in line with the amyloid cascade hypothesis and the A/T/N framework that genetic risk in Alzheimer’s disease is disease-stage dependent. That is, some genetic risk factors will aid the transition from A- to A+, while other, distinct genetic risk factors will increase the risk to transition from T- to T+.

In this work we explore whether genetic vulnerability to Alzheimer’s disease varies with disease stage. Using longitudinal data from the Alzheimer’s disease neuroimaging imitative (ADNI) and survival analysis we show that *APOE* contributes to progression from A-T- to A+T-, but only marginally from A+T- to A+T+. Conversely, polygenic risk contributes to the progression from A+T- to A+T+, but not from A-T- to A+T-.

## Materials and methods

### Data

Data used in the preparation of this article were obtained from the ADNI database (http://adni.loni.usc.edu). The ADNI was launched in 2003 as a public–private partnership, led by Principal Investigator Michael W. Weiner, MD. The primary goal of the ADNI has been to test whether serial MRI, PET, other biological markers, and clinical and neuropsychological assessment can be combined to measure the progression of mild cognitive impairment (MCI) and early Alzheimer’s disease. For up-to-date information, see www.adni-info.org. ADNI study data were accessed through the R-package ADNIMERGE (accessed: 20^th^ July 2023).

### Preparation of genetic data

The genetic data preparation followed the procedure described in Altmann et *al.*^32^. The additional genetic data contributed by the ADNI-3 cohort was integrated with the existing data using the same processing pipeline. Briefly, single nucleotide polymorphism (SNP) genotyping data were available for *n* = 2001 subjects across ADNI phases 1,2, GO and 3. Genotyping was conducted using four different platforms: Human610-Quad, HumanOmniExpress, Omni 2.5 M and Illumina Infinium Global Screening Array v2 (Illumina)^38^. Prior to imputation, we applied subject-level quality control (QC) steps based on call rate (10% cutoff) and concordance between chip-inferred sex and self-reported sex separately for the four genotyping arrays, all subjects were retained. On SNP-level, we conducted standard QC steps ensuring compatibility with the reference panel used for imputation [strand consistency, allele names, position, Ref/Alt assignments and minor allele frequency (MAF) discrepancy (0.2 cutoff)]. Imputation was carried out using the Sanger Imputation Server (https://imputation.sanger.ac.uk/) with SHAPEIT for phasing^39^, Positional Burrows-Wheeler Transform^40^ for imputation and the entire Haplotype Reference Consortium (release 1.1) reference panel^41^. Data from the four different genotyping platforms were imputed separately. As part of post-imputation QC, multi-allelic variants and SNPs with imputation INFO score <0.3 were removed and genotype calls with posterior probability <0.9 were set to missing (i.e., hard called). Following the initial QC, genotypes from the four platforms were merged. Additional information on the imputation and QC process is detailed in Scelsi *et al.*^28^ and https://rpubs.com/maffleur/452627. Using the merged data, we retained SNPs with MAF ≥1% and genotyping rate >0.9.

SNPweights^42^ was used to infer genetic ancestry from genotyped SNPs. The reference panel comprised Central European, Yoruba Africans and East Asian samples from HapMap 3^43^ and native Americans from Reich *et al.*^44^. For this study, only participants with predicted central European ancestry of 80% or more were retained (*n*=1851). Next, using the imputed and merged data genetic relatedness between central European participants was computed. First, the SNP content was restricted to SNPs with MAF ≥5% and linkage disequilibrium (LD)- pruning was carried out in PLINK v1.9 (–indep-pairwise 1000 50 0.1). The genetic relatedness matrix was computed using the remaining autosomal SNPs and the dataset was trimmed to remove subjects with relatedness >0.1 (–rel-cutoff 0.1), leading to *n*=1833 unrelated participants.

### Definition of genetic risk

In this study, we focused on two sources of genetic risk: (1) the risk conferred through the *APOE* gene based on the genetic markers for APOE-ε2 and APOE-ε4 and (2) the polygenic risk conferred by the remaining genome. As described previously^32^, PRSs were computed using the software PRSice v2.1.9^45^. As base GWAS the stage 1 results of the Alzheimer’s disease GWAS featuring a clinically defined Alzheimer’s disease phenotype was used^18^. For PRS computation, SNPs with MAF ≥5% were considered and SNPs were selected using LD-clumping (1000 KB, R2 of 0.1 and P-value threshold of 1.0) within the ADNI cohort, missing SNPs were simply ignored on subject level (using the setting–missing SET_ZERO) and the *APOE* region was excluded (hg19 coordinates chr19 from 44,400,000 to 46,500,000). For this study, we used only the P-value cutoff of 1.0e-8 to build the PRS (**Supplementary Table 1**). PRSs were computed for all ADNI participants with genome-wide genotyping data. Of the remaining subjects, *n*=417 ADNI-1 participants who contributed to the Alzheimer’s disease GWAS^18^ were excluded from the analysis to ensure independence between training and application dataset for PRS. Thus, *n*=1416 unrelated participants with central European ancestry were eligible for inclusion into the study.

### Definition of amyloid status

For this project we relied on two modalities to define amyloid status: Amyloid-β PET using the ^18^F-florbetapir (FBP) and the ^18^F-florbetaben (FBB) PET tracer, and CSF measures of amyloid-β (1-42) using the Roche Elecsys® immunoassay. We used processed data by ADNI for both modalities. Detailed information on the PET processing is available elsewhere (http://adni-info.org)^46^; and information on CSF amyloid-β(1-42) processing is detailed in^47,48^. For CSF amyloid-β(1-42) we used the cutoff of 880 pg/mL^48^ and for amyloid-β PET we used the tracer-specific SUVR (whole cerebellum reference) cutoffs of 1.11 and 1.08 for FBP and FBB^49^, respectively.

A participant’s visit was labelled as A+ if either the PET result or the CSF result indicated a positive amyloid finding (i.e., in cases where the results were discordant, the visit would be labelled as A+). Visits with only negative amyloid findings were labeled as A- and visits without any information on amyloid (i.e., neither a PET nor a CSF result) were labelled as ‘Amyloid missing’.

### Definition of tau status

In keeping with our definition of amyloid positivity we used available data from CSF and PET imaging to define the tau status. More precisely, we used the Phospho-Tau(181P) CSF Roche Elecsys® immunoassay and PET imaging using the ^18^F-Flortaucipir tracer. Details on the processing are available elsewhere (http://adni-info.org)^47,48,50^. For CSF we used phosphorylated 181P tau (pTau) with a cutoff of 34.61 pg/mL and for tau PET we used the cutoff of 1.42 in the meta temporal ROI comprising amygdala, entorhinal cortex, fusiform gyrus, inferior and middle temporal gyri^51^ when normalized to the inferior cerebellar gray matter^52^. Both cut-offs were data-driven: (1) the tau PET cutoff corresponds to a z-score of 2.0 in the cognitively normal participants in ADNI (*n*=506) and is close to the ‘high tau’ cutoff of 1.43 defined in by Jack Jr *et al.* ^53^; (2) the pTau cutoff was set to maximize the Youden’s index between CSF pTau and tau PET (at the 1.42 cutoff) in (*n*=502) ADNI participants with concurrent CSF and PET measurements. The same labeling scheme as for amyloid was applied: a visit was labelled as T+ if either the CSF or the PET indicated a positive finding, T- if there were only negative tau findings and ‘Tau missing’ if neither data on CSF pTau nor on tau PET were available.

### Statistical modeling

We used cox-proportional hazards models to investigate the genetic contribution of progressing from (1) A-T- to A+T- and from (2) A+T- to A+T+. For (1) we included every eligible participant with genetic data and who was A-T- based on their biomarker results as described above. This earliest A-T- visit was considered the ‘start’ visit (i.e., the status of previous visits was ignored). A subject was considered a converter when the available biomarkers indicated A+T- at a later visit. The time of the first A+T- biomarker finding after their initial A-T- visit was used as the conversion time. For non-converters we recorded the last visit where both amyloid and tau biomarker information was available to define the maximum follow-up time. Similarly, for (2) we included every participant with genetic data and who was A+T-. This first A+T- visit was considered the ‘start’ visit. A subject was considered a converter when the available biomarkers indicated A+T+ at a later visit. The time of the first A+T+ biomarker finding after their initial A+T- visit was used as the conversion time. As before, for non-converters we recorded the last visit where amyloid and tau biomarker information was available as their last point of contact.

For the analyses, the cox-proportional hazards model included age at the inclusion visit (i.e., either the A-T- or the A+T- visit), sex, and education. As variables of interest, we further included genetic variables for PRS as well as allele counts of APOE-e2 and APOE-e4. The proportional hazards assumption was tested for each covariate and for the global model using statistical tests and graphical diagnostics based on scaled Schoenfeld residuals. As a measure of overall model performance, the concordance index (C-index) was computed for the (i) full models (ii) models without *APOE*, (iii) models without PRS, and (iv) models without any genetics. Statistical tests were carried out in R (v 4.1.0) using the `survival` (v 3.5.5), `survminer` (v 0.4.9), and `rms` (v 6.8.0) packages.

### Sensitivity analyses

In addition to the two main analyses, we conducted a series of sensitivity analyses addressing the conversion definition, biomarker cutoffs, biomarker source, and polygenic score source.

### Conversion definition

To maximize the available data, we relaxed the requirement for both amyloid and tau biomarker to be available at the same visit to define conversion. As before, both biomarkers at the same time were required to define the ‘start’ visit as either A-T- or A+T-. However, for defining progression from A-T- or A+T-, a single A+ or T+ visit was sufficient, respectively.

### Biomarker cutoffs

We varied the tau PET cutoff from 1.0 to 3.0 standard deviations above the mean in the cognitively normal ADNI participants. Notably, the lowest cutoff resulted in 1.31 and was close to the neuropathologically defined cutoff of 1.29 by Lowe *et al.*^54^. The pTau cutoffs were adjusted accordingly to maximize Youden’s index between CSF pTau and tau PET.

### Biomarker source

To maximize data and follow-up time, the main analyses combined data from two biomarker sources: CSF and PET. Additional sensitivity analyses relied exclusively on either CSF biomarkers or PET biomarkers. For this analysis pTau cutoffs were varied in the range from 22 to 31, covering the values of 24.25 and 29.19, which were found to indicate tau PET positivity in Braak III/IV and Braak V/VI regions, respectively^55^. Tau PET cutoffs were varied, as before, from 1.0 to 3.0 standard deviations above the mean in cognitively normal participants (i.e., in the same range as the main analysis).

### Polygenic Score source

To include more SNPs into the PRS, we explored the summary statistics on Alzheimer’s Disease and related dementias by Bellenguez *et al.*^21^. We followed the same PRS pipeline as above and applied a p-value threshold of 5.0e-08 (i.e., genome-wide significant) leading to 77 included SNPs (**Supplementary Table 2**).

## Results

Out of 16,401 visits recorded in ADNI, 3789 (23.1%) visits had both amyloid and tau biomarker data available. Concordance between PET- and CSF-based assessment was 80% and 81% for amyloid and tau, respectively. For both survival models we identified around 300 subjects in the ADNI database with both biomarkers available (**Table 1**): 312 individuals were A-T- of whom 65 converted to A+T-. The mean age of the A-T- group was 71.3 (6.65) years and there was an almost equal number of males and females (49.4% females). *n*=290 individuals were A+T- at any stage, of whom 45 converted to A+T+. The mean age of the A+T- group was 73.2 (6.9), significantly older than the A-T- group (unpaired t-test: *T*=3.50, *P*<0.001). The fraction of females in that cohort was lower compared to the A-T- group (43.8% females), but not at a significant level (chi-square-test; chi-square=1.65; df=1; *P*=0.19). The distribution of APOE-e4 alleles (chi-square test; chi-square= 81.1; df=2; *P*=2.4e-18) and APOE-e2 alleles (Fisher’s exact test; *P*= 0.0004) differed significantly between A-T- and A+T. There were more APOE-e2 carriers and fewer APOE-e4 carriers in the A-T- group than in the A+T- group. The PRS did not differ between the A-T- and A+T- groups (two-sided t-test; t=0.84; df=600; *P*=0.39). The cohort with the relaxed conversion criterion showed comparable characteristics (**Supplementary Table 3**).

**Table 1:** Demographics.

|  | **A-T-** |  |  | **A+T-** |  |  |
| --- | --- | --- | --- | --- | --- | --- |
|  | **Total** | **Stable** | **Converter** | **Total** | **Stable** | **Converter** |
| *n* | 312 | 247 | 65 | 290 | 245 | 45 |
| Age, y (sd) | 71.3 (6.65) | 71.0 (6.47) | 72.2 (7.25) | 73.2 (6.9) | 73.2 (6.9) | 73.1 (6.8) |
| Sex (%female) | 49.4 | 48.6 | 52.3 | 43.8 | 43.3 | 46.7 |
| DX (CN/MCI/AD) | 195/113/4 | 150/94/3 | 45/19/1 | 118/153/19 | 100/128/17 | 18/25/2 |
| Education, y (sd) | 16.7 (2.5) | 16.6 (2.6) | 17.3 (2.2) | 16.4 (2.6) | 16.6 (2.7) | 15.5 (2.4) |
| Years Follow-up, y (sd) | 5.0 (3.1) | 4.7 (3.0) | 6.4 (3.4) | 4.1 (2.6) | 3.8 (2.6) | 5.6 (2.3) |
| Time to event, y (sd) | n/a | n/a | 4.5 (2.7) | n/a | n/a | 4.4 (2.5) |
| APOE-e4 (0/1/2) | 252/57/3 | 207/40/0 | 45/17/3 | 137/119/34 | 121/99/25 | 16/20/9 |
| APOE-e2 (0/1/2) | 265/46/1 | 205/41/1 | 60/5/0 | 273/17/0 | 230/15/0 | 43/2/0 |
| PRS (sd) | 0.013 (0.012) | 0.013 (0.012) | 0.013 (0.011) | 0.013 (0.013) | 0.012 (0.013) | 0.020 (0.011) |

DX = diagnosis (cognitively normal [CN]; mild cognitive impairment [MCI]; Alzheimer's Disease (AD)); PRS = Polygenic Risk Score

### APOE-e4 influences progression from A-T- to A+T-

The survival analysis showed a significant contribution by APOE-e4 allele count (HR=2.88; 1.70-4.89; *P*=8.7e-05) but no significant contribution by PRS (HR=1.09; 0.84-1.42; *P*=0.53) (**Fig. 1**). The APOE-e2 allele count directionally favoured a protective effect, but this was not significant (HR=0.73; 0.28-1.86; *P*=0.51). The Cox proportional hazards assumption held for this model (Global Schoenfeld test *P*=0.24) (**Supplementary Fig. 1**). The C-indices align with this pattern: full model (C=0.612), no *APOE* (C=0.525), no PRS (C=0.611), no genetics (C=0.531). This pattern of associations of *APOE* and PRS with progression was largely independent of the tau PET and pTau thresholds (**Supplementary Table 4**). Furthermore, using the more relaxed conversion criteria (i.e., confirmed A+ status was sufficient instead of a confirmation of A+T-) yielded more conversions (85 instead of 65) but qualitatively the same result (**Supplementary Fig. 2**), i.e., a significant contribution by APOE-e4 allele count (HR=3.34; 2.14-5.22; *P*=1.0e=07) but not by PRS (HR=1.06; 0.84-1.34; *P*=0.61).

**Figure 1:**
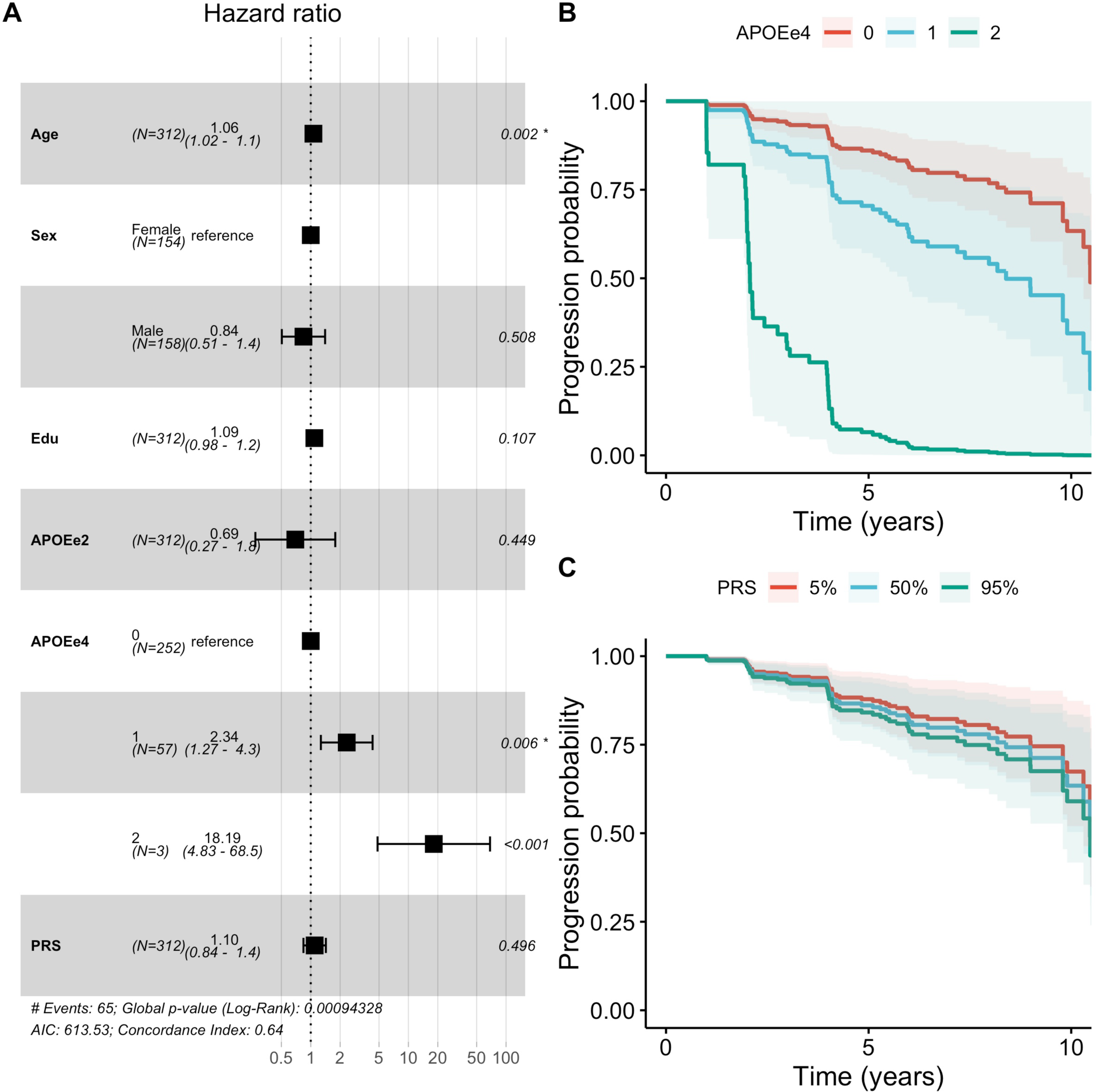
Hazard ratios for the conversion from A-T- to A+T-. (A) Forest plot depicting the Hazards Ratios (HR) for all covariates in the model. (B) Estimated survival curves stratified by APOEe4 genotype. (C) Estimated survival curves stratified by PRS percentile (5%, 50%, 95%). Edu = Years of Educations; APOEe2 = number of APOE2 alleles, APOEe4 = number of APOE4 alleles; PRS = polygenic risk score, scaled to zero mean and unit standard deviation.

### Polygenic risk affects the progression from A+T- to A+T+

The survival analysis showed a marginally significant contribution by APOE-e4 burden (HR=1.62; 1.05-2.51; *P*=0.031), which was mainly driven by APOE-e4 homozygotes (HR=2.58; 1.05-6.35; *P*=0.039) rather than APOE-e4 heterozygotes (HR=1.74; 0.87-3.49; *P*=0.12) (**Fig. 2**). Furthermore, there was a significant contribution by PRS (HR=1.72; 1.27- 2.36; *P*=0.00057). The APOE-e2 allele count again was directionally consistent with a protective effect, but this was not significant (HR=0.41; 0.09-1.78; *P*=0.23). The Cox proportional hazards assumption held for this model (**Supplementary Fig. 3**). The C-indices drop marginally when either *APOE* or PRS are removed from the model: full model (C=0.657), no *APOE* (C=0.634), no PRS (C=0.615), no genetics (C=0.549). Moreover, the association pattern of *APOE* and PRS with progression was largely independent of the tau PET and pTau cutoffs (**Supplementary Table 4**). In addition, education showed a marginally protective association with conversion to A+T+ (HR=0.89; 0.80-0.99; *P*=0.039). Applying the more relaxed conversion criteria (i.e., confirmed T+ status was sufficient instead of a confirmation of A+T+) yielded more conversions (51 instead of 45) but qualitatively the same result (**Supplementary Figure 5**): a significant contribution by PRS (HR=1.62; 1.22-2.17; *P*=0.001) and a marginal contribution by APOE-e4 allele burden (HR=1.56; 1.04-2.35; *P*=0.031).

**Figure 2:**
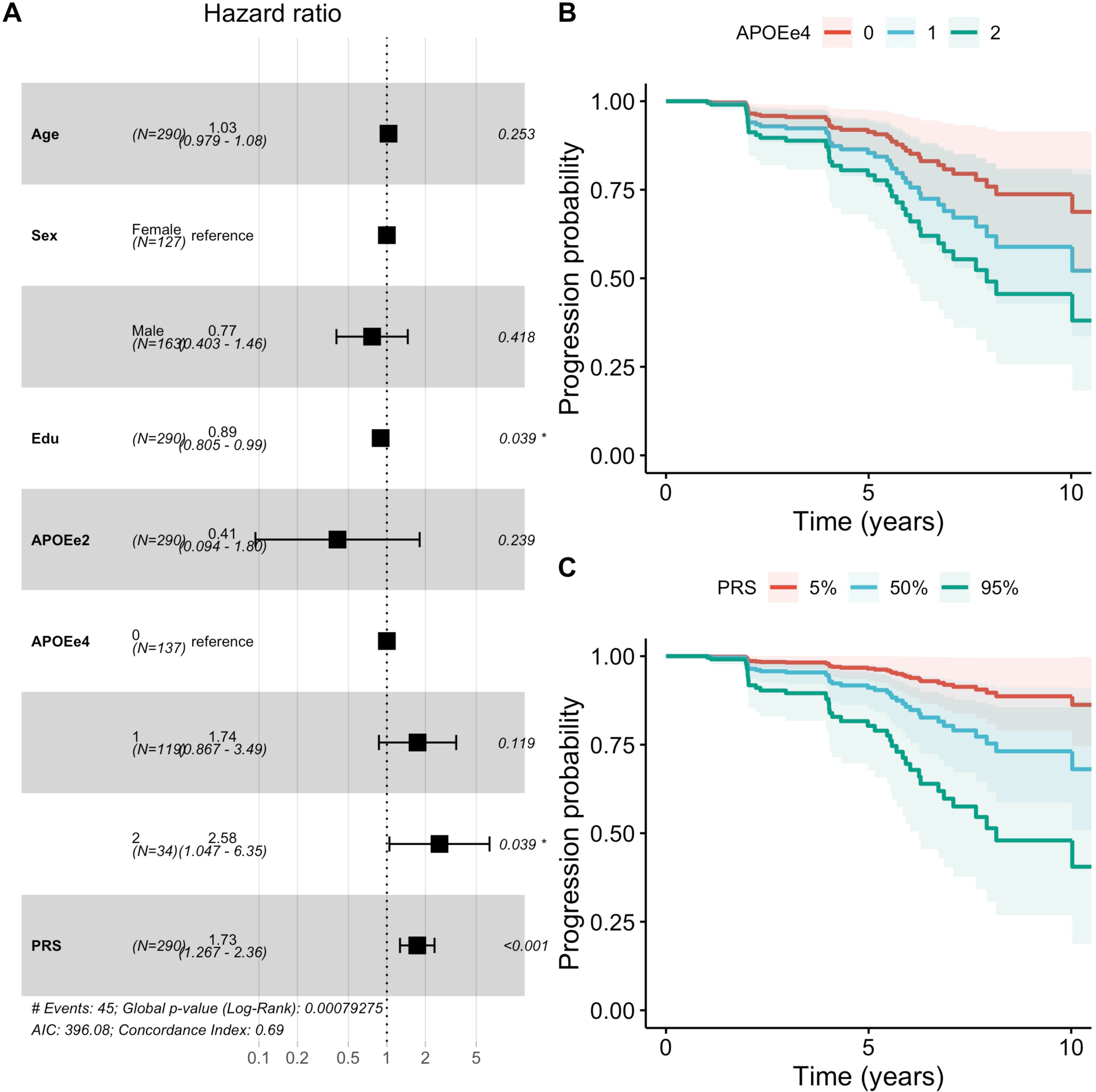
Hazard ratios for the conversion from A+T- to A+T+. (A) Forest plot depicting the Hazards Ratios (HR) for all covariates in the model. (B) Estimated survival curves stratified by APOEe4 genotype. (C) Estimated survival curves stratified by PRS percentile (5%, 50%, 95%). Edu = Years of Educations; APOEe2 = number of APOE2 alleles, APOEe4 = number of APOE4 alleles; PRS = polygenic risk score, scaled to zero mean and unit standard deviation.

### Results are independent of PRS source

Using an the alternative PRS with 77 SNPs led to the same observation of a significant effect by APOE-e4 burden on A-T- to A+T- conversion (HR=2.84; 1.68-4.82; *P*=0.0001) and a lack of contribution by the PRS (HR=0.97; 0.77-1.2; *P*=0.80; **Supplementary Fig. 5**A). Conversely, for A+T- to A+T+ conversion, the contribution by APOE-e4 was reduced (HR=1.71; 1.09-2.68; *P*=0.019), where the PRS exhibited a strong contribution (HR=1.59; 1.19-2.13; *P*=0.00163; **Supplementary Fig. 5**B).

### Results are independent of biomarker source

The main analysis combined different biomarker sources to maximize the available data and the observation time for the conversion analysis. Just relying on PET biomarkers alone, only 7.5% of visits (1237 of 16,401) had both biomarkers and it resulted in shorter observation times for A-T- to A+T- conversion (3.62 (sd=1.07) years) and A+T- to A+T+ conversion (2.94 (sd=1.13) years) compared to the main analysis. Relying solely on CSF biomarkers, 19.2% of visits (3155 of 16,401) had both biomarkers. Overall, this led to shorter observation time with respect to the main analysis for A-T- to A+T- conversion (3.94 (sd=2.7) years) and A+T- to A+T+ conversions (3.6 (sd=2.38) years). Furthermore, relying on a single source of biomarkers led to reduced sample sizes (from around 300 in the main analyses to 70-250 in the sensitivity analyses) and observed conversions (**Supplementary Table 5** and **Supplementary Table 6**). However, despite the reduced statistical power these sensitivity analyses confirmed the pattern observed in the main analysis: APOE-e4 contributed mainly to the A-T- to A+T- conversion, while PRS contributed to the A+T- to A+T+ conversion **(Fig. 3, Supplementary Table 5, Supplementary Table 6).**

**Figure 3:**
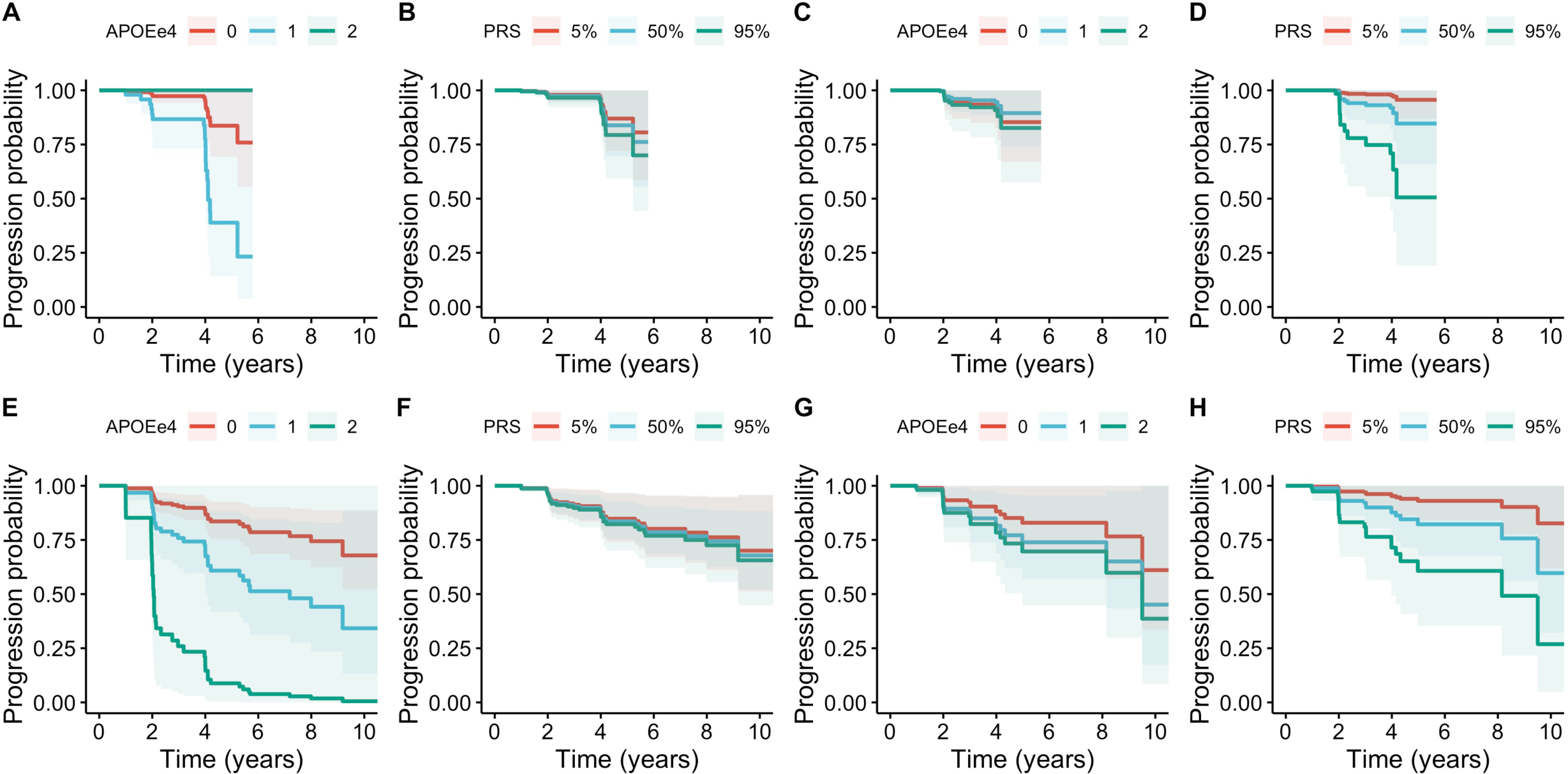
Survival curves for PET only and CSF only analyses. The top row (A-D) is based on results from AT(N) definitions based exclusively on PET biomarkers (amyloid and tau) with a cutoff of 1.45 for tau PET; the bottom row (E-H) relied on CSF biomarkers (ABETA42 and pTAU) with a cutoff of 26 for pTau. The two left columns (A, B, E, F) display the progression probability from A-T- to A+T- stratified by APOEe4 genotype and PRS quantile (5%, 50%, 95%). The two right columns (C, D, G, H) display the progression probability from A+T- to A+T+.

## Discussion

This longitudinal survival analysis demonstrated that APOE-e4 plays an important role for the progression from A-T- to A+T-, but *APOE* is only of marginal importance in A+T- participants who progress to A+T+. Conversely, polygenic risk for AD exhibited the inverse pattern: there was no contribution to the progression from A-T- to A+T-, but a significant contribution to faster progression from A+T- to A+T+. This held true for an alternative PRS defined using a different genetic study and involving a larger number of genetic loci. Notably, when assessing covariates, a differential effect of years of education was observed: higher education had no effect (**Fig. 1**) or was marginally harmful (**Supplementary Fig. 2**) for the progression from A- T- to A+T-, but was protective for the conversion from A+T- to A+T+ (**Fig. 2**). The bisection of the genetic risk by disease stage was largely independent of the applied biomarker cutoffs. Moreover, relying on only a single source of biomarkers for defining stage and conversion confirmed the findings of the main analysis despite reduced sample size and observation time.

The finding of a stronger effect of APOE-e4 earlier in the disease progress may explain the observation of stronger genetic effect of APOE-e4 on Alzheimer’s disease in the group of 60-80 year-old people compared to people 80 years and older^56–58^. Amyloid deposition occurs 10+ years before other Alzheimer’s disease processes^2^ and APOE-e4 is the strongest common genetic risk factor for amyloid deposition, then it would be expected for APOE-e4 to exert its maximum effect in younger people. Still, *APOE* remains the strongest risk factor in individuals 80 years and older^57^. Thus, the age-dependent heterogeneity of *APOE* is likely compounded by a survivor bias: individuals with a very late onset despite carrying APOE-e4 may harbor protective variants^59^ such as KLOTHO-VS, where a protective effect on amyloid deposition and Alzheimer’s disease was only observed in 60-80 year-olds, but not in the 80+ group^56,60^.

The findings from this longitudinal analysis are also in line with previous reports in the ADNI cohort of *APOE* and polygenic risk on amyloid and tau. For instance, cross-sectional amyloid biomarkers in the CSF and in the brain were mainly driven by *APOE*, while cross-sectional CSF tTau and pTau measurements were associated with PRS beyond the *APOE* locus^32^. Moreover, *APOE* was found to predict amyloid status while polygenic risk for Alzheimer’s disease improved predictions of diagnosis as well as clinical progression from MCI to Alzheimer’s disease above *APOE* alone^35^. These observations extend to plasma markers of tau pathology: PRS (that excluded the *APOE* region) were found to be only associated with plasma p-tau181 in A+ participants^61^. This association between polygenic risk (beyond *APOE)* and CSF tau biomarkers rather than with amyloid and neurodegeneration was also observed outside of the ADNI study^62^. Polygenic risk (beyond *APOE*) was associated with non-amyloid endophenotypes in a large cohort of people with MCI. This suggests that these variants are more closely linked with neuronal degeneration than with Alzheimer’s disease-related amyloid pathology^63^. All these previous studies made the connection between existing amyloid pathology and correlations between polygenic risk and tau pathology using cross-sectional study designs. In a recent longitudinal study of tau PET in the ADNI cohort, higher polygenic risk for Alzheimer’s disease was associated with accelerated increase in tau signal in brain and this effect was modulated by amyloid pathology: A+ participants showed a stronger effect of PRS on tau accumulation^64^. Our longitudinal analysis, which combined CSF and PET data to maximize the sample size, confirms these observations and indicates that polygenic Alzheimer’s disease risk (outside the *APOE* region) contributes to tau pathology in A+ participants, but has no meaningful contribution in A- participants. The genetic architecture of Alzheimer’s disease, as captured by the PRS, involves multiple different pathways, mainly β-amyloid processing, tau, immunity, and lipid processing^18,21^. The employed PRS covers three genes that have been associated with tau binding: *BIN1*, *CLU* and *PICALM*. *BIN1* mediates Alzheimer’s disease risk by modulating tau pathology^65^, and *BIN1* risk variants increase tau PET (but not amyloid PET)^66^ in an amyloid-dependent fashion^67^. Our observations of an amyloid-dependent effect of the PRS align with these earlier single gene studies. While there are currently no mechanistic analyses that explain the amyloid-dependent effect of *BIN1* on tau pathology, recent data from animal models in Alzheimer’s disease suggest a state-dependent effect of genetic risk factors related to microglia: Deletion of *Trem2* in mouse models exacerbated tau accumulation and spreading leading to brain atrophy, but only in the presence of existing β-amyloid pathology^68^. Along the same line, physical contact between microglia and plaques as well as a functioning *TREM2* gene are necessary for the appropriate microglia response to amyloid pathology^69^. Thus, defects in *TREM2* can only contribute to neurodegeneration once amyloid pathology has been established. Consequently, other genes contributing to the PRS may also exert their effect in an amyloid-dependent fashion. Our longitudinal analysis presented here is the first to support such a state-dependent genetic risk model in humans, and further fine-grained examination of how the pathways involved in the PRS contribute to sequential disease progression are needed.

The partition of Alzheimer’s disease genetic risk into *APOE*-related and polygenic risk beyond *APOE* is a simplification in this analysis. Recent works have linked established Alzheimer’s disease risk loci outside *APOE*, such as *CR1*, to amyloid biomarker levels^23^. Conversely, studies of biomarker levels of tau repeatedly highlight the *APOE* locus^23,24^. However, if being A+ were a pre-requisite to exhibit pathological accumulation of tau, then the strong genetic association with *APOE* in these GWASs would merely reflect the necessary condition rather than a genuine direct molecular process that affects tau levels. The strong dependency of tau levels on established amyloid pathology is supported by mediation analyses in recent cross-sectional^62^ and longitudinal^64^ studies. Moreover, the known Alzheimer’s disease genetic risk variants are contributing differently to molecular pathways^18,21,22^, where each pathway in turn will exercise differential effects on the AD biomarkers including markers for vascular pathology^37^. Therefore, a pathway PRS only comprising genes associated with the regulation of the amyloid precursor protein catabolic process (e.g., Gene Ontology term GO:1902991) may contribute significantly to the conversion from A-T- to A+T-. Likewise, a pathway PRS using only genes known to bind the tau protein (GO:0048156) may exhibit an even stronger association with A+T- to A+T+ conversion than the general PRS employed here.

In addition to the cascading effect of genetic risk in Alzheimer’s disease, we also observed a stage-dependent effect of non-genetic risk factors. Education has been shown to have a protective effect against dementia^70^: here we show that higher rates of education do not influence transition to amyloid positivity, but do slow progression from A+T- to A+T+. Consequently, other non-genetic risk factors may show a similarly state-dependent effect on the pathological pathway from A-T- to A+T+ and further neurodegeneration.

The study has several limitations. Firstly, the current analysis was limited to just two biomarkers in Alzheimer’s disease: β-amyloid and tau. It would be desirable to include neurodegeneration (N; of the ATN framework) or potentially more fine-grade staging from advanced data-driven disease progression modeling^7^. However, at this point adding further stages would reduce the available sample size. Secondly, we partitioned the genetic risk in Alzheimer’s disease into two components: *APOE* and other top variants combined into a single polygenic risk score. Further work should explore a more fine-grained partition of the polygenic risk into individual SNPs or into pathway-PRS^37^. Thirdly, the study population was of central European ancestry, therefore it is unclear whether the findings would generalize to other genetic backgrounds. Finally, although the ADNI cohort is a large cohort, the number of subjects who were eligible for our analysis was reduced due to the requirement of concordant and longitudinal recordings of multiple biomarkers as well as genetics. The available sample size may have limited statistical power to render the estimated hazards ratios significant in some settings. However, the two conversion analyses were based on similar sample sizes (around 300 participants), thus allowing us to make a relative comparison between the genetic effect (of *APOE* or PRS) under two different, biomarker defined, disease stages. Moreover, uncertainty of the estimated effects may also be increased due to disease heterogeneity in Alzheimer’s disease^71–74^, which is likely underpinned by differences in genetic architecture. Thus, analyses in further large longitudinal cohorts are required to confirm the observation of stage-dependent genetic vulnerability in Alzheimer’s disease and to uncover more fine-grained associations with Alzheimer’s disease subtypes.

In this work we demonstrated, in a simplified setting, that genetic risk for late onset Alzheimer’s disease unfolds in a disease stage-dependent fashion. A better understanding of the interplay between disease stage and genetic risk can lead to a better understanding of the molecular processes leading to Alzheimer’s disease as well as opening therapeutic windows for targeted interventions and personalized approaches to dementia prevention.

## Data Availability Statement

Data used in the preparation of this article were obtained from the ADNI database (http://adni.loni.usc.edu) and are freely available after registration. Analysis scripts are available at https://github.com/andrealtmann/cascading_genetic_risk.

## Supplementary material

Supplementary material is available at *Brain* online.

## Data Availability

Data used in the preparation of this article were obtained from the ADNI database (http://adni.loni.usc.edu) and are freely available after registration.

## Supplementary Material

**Supplementary Figure 1:**
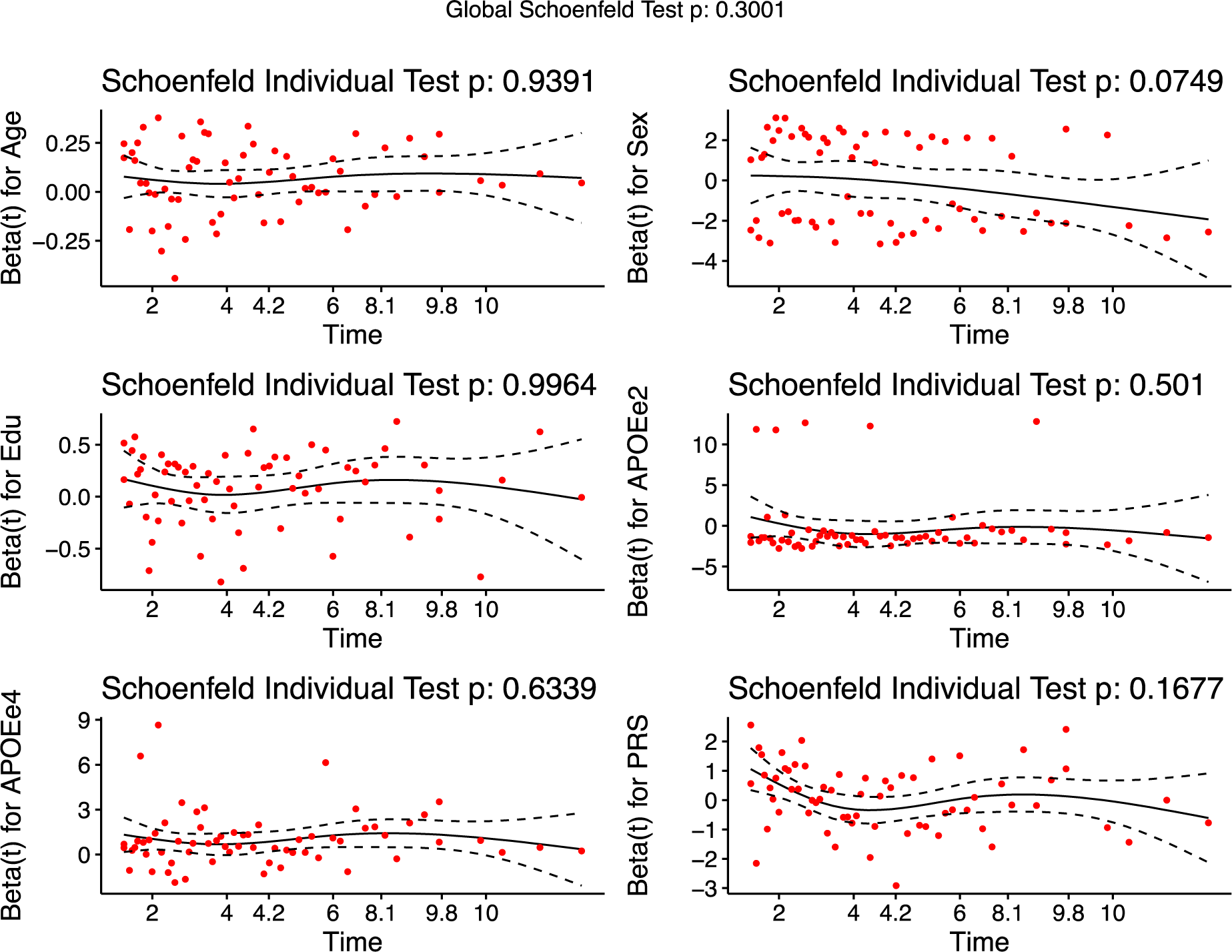
Testing the Cox Proportional Hazards assumption for the Amyloid model. Each panel depicts the scaled Schoenfeld residuals for one covariate. The title of each panel states the p-value of the individual Schoenfeld tests. The title of the plot states the p-value of the global Schoenfeld test. None of the p-values are significant (i.e., *P*<0.05), therefore the Cox Proportional Hazards assumption holds.

**Supplementary Figure 2:**
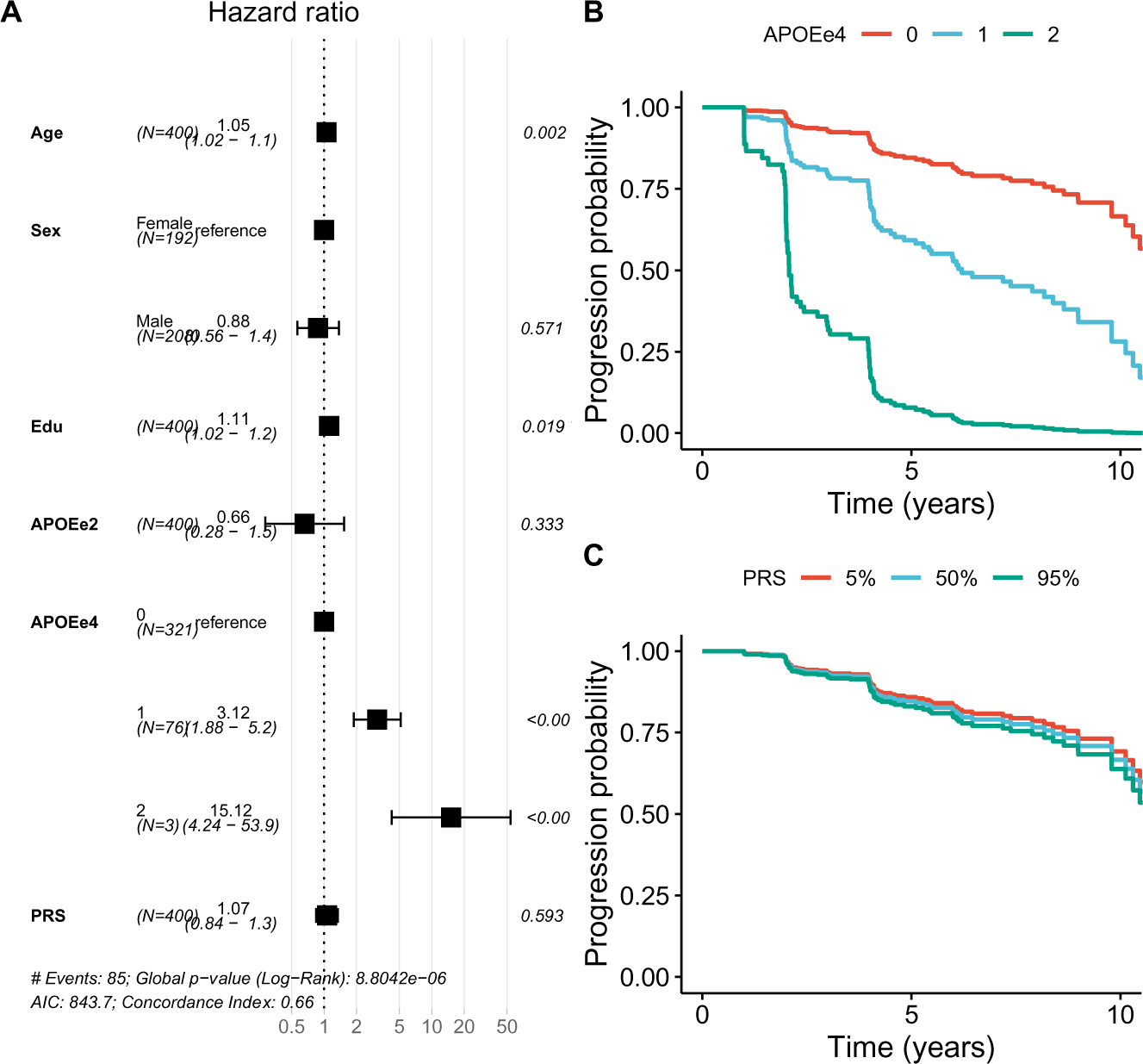
Hazard ratios for the conversion from A-T- to A+. (A) Forest plot depicting the Hazards Ratios (HR) for all covariates in the model. (B) Estimated survival curves stratified by APOEe4 genotype. (C) Estimated survival curves stratified by PRS percentile (5%, 50%, 95%). Edu = Years of Educations; APOEe2 = number of APOE2 alleles, APOEe4 = number of APOE4 alleles; PRS = polygenic risk score, scaled to zero mean and unit standard deviation.

**Supplementary Figure 3:**
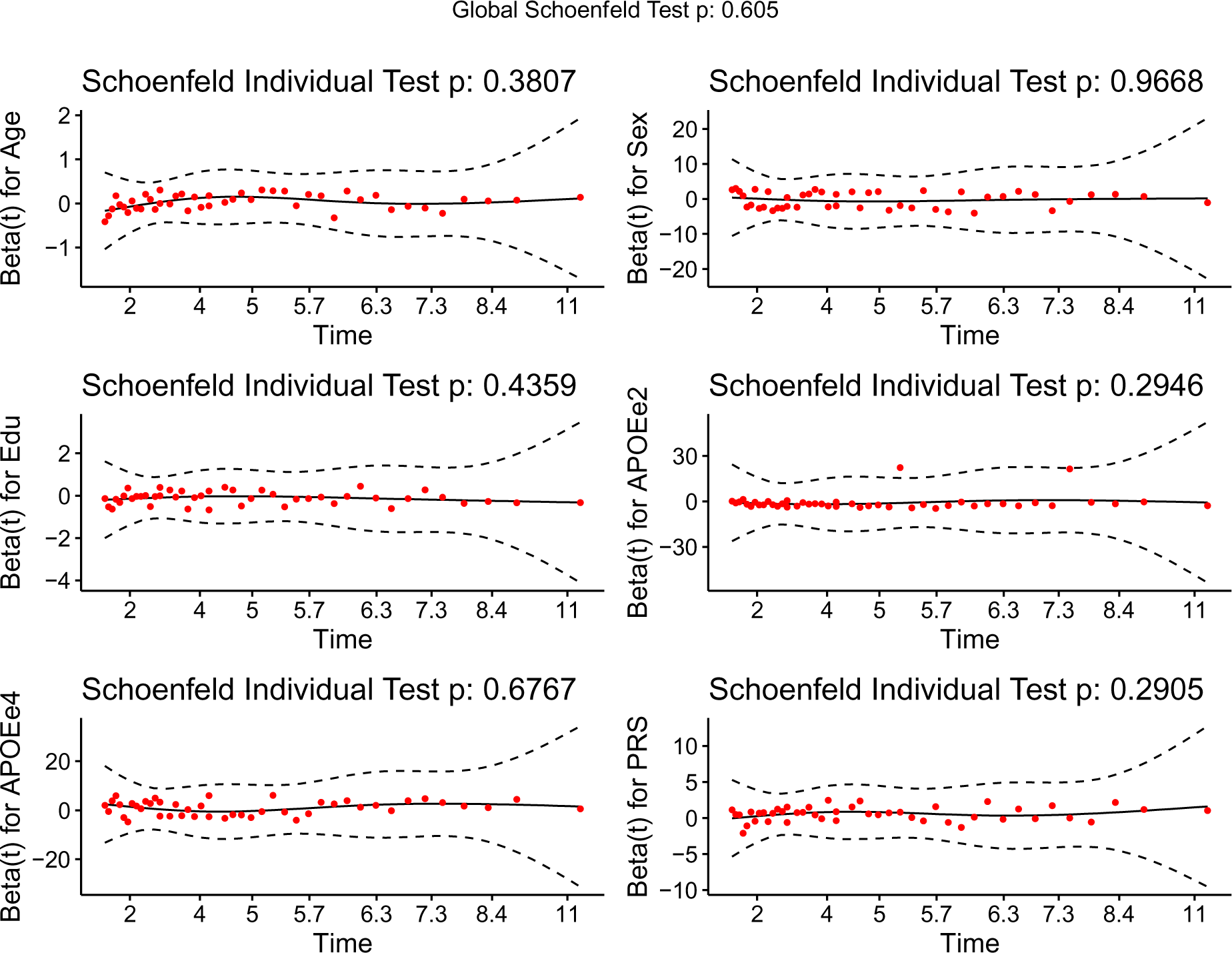
Testing the Cox Proportional Hazards assumption for the Tau model. Each panel depicts the scaled Schoenfeld residuals for one covariate. The title of each panel states the p-value of the individual Schoenfeld tests. The title of the plot states the p-value of the global Schoenfeld test. None of the p-values are significant (i.e., *P*<0.05), therefore the Cox Proportional Hazards assumption holds.

**Supplementary Figure 4:**
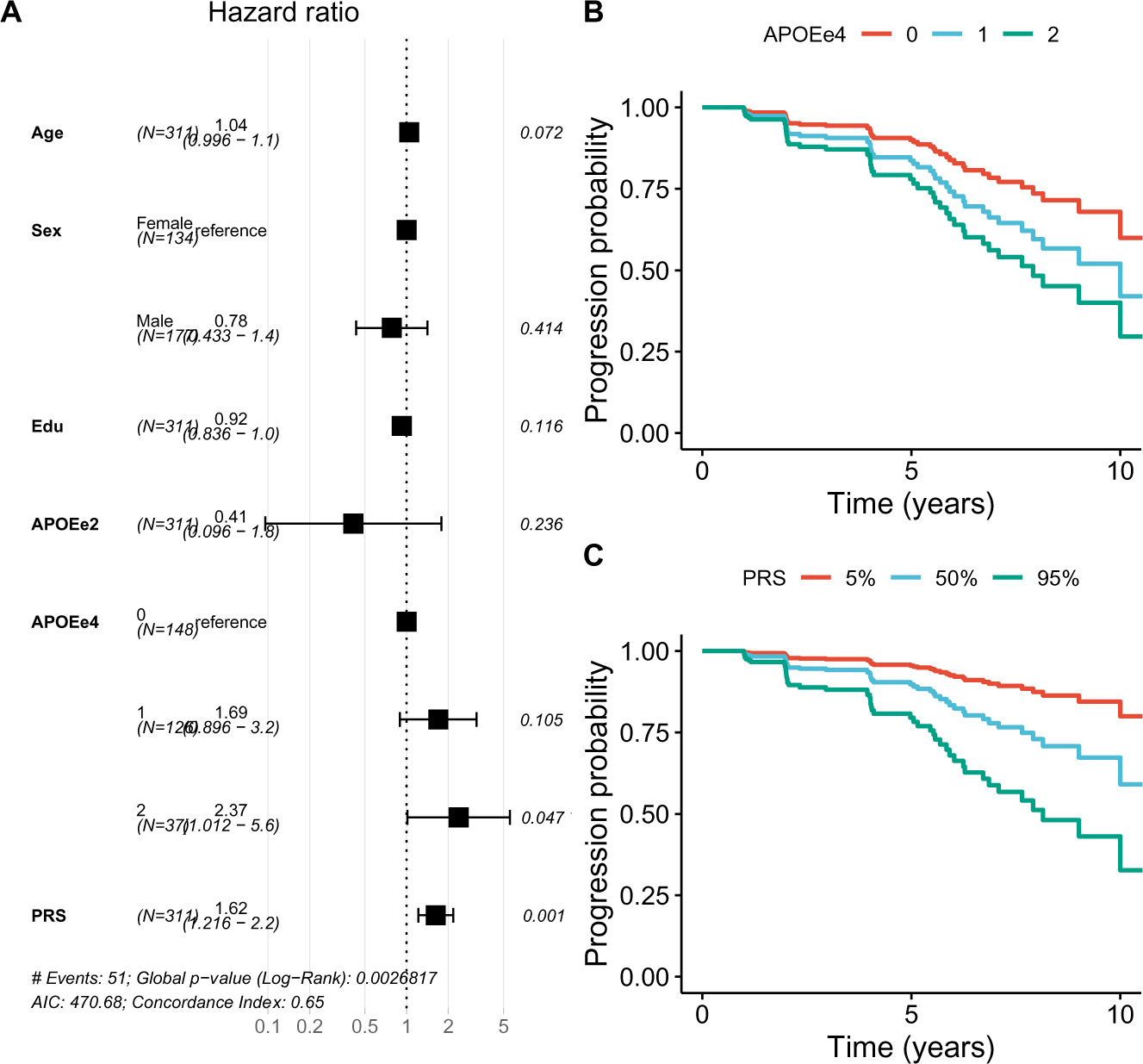
Hazard ratios for the conversion from A+T- to T+. (A) Forest plot depicting the Hazards Ratios (HR) for all covariates in the model. (B) Estimated survival curves stratified by APOEe4 genotype. (C) Estimated survival curves stratified by PRS percentile (5%, 50%, 95%). Edu = Years of Educations; APOEe2 = number of APOE2 alleles, APOEe4 = number of APOE4 alleles; PRS = polygenic risk score, scaled to zero mean and unit standard deviation.

**Supplementary Figure 5:**
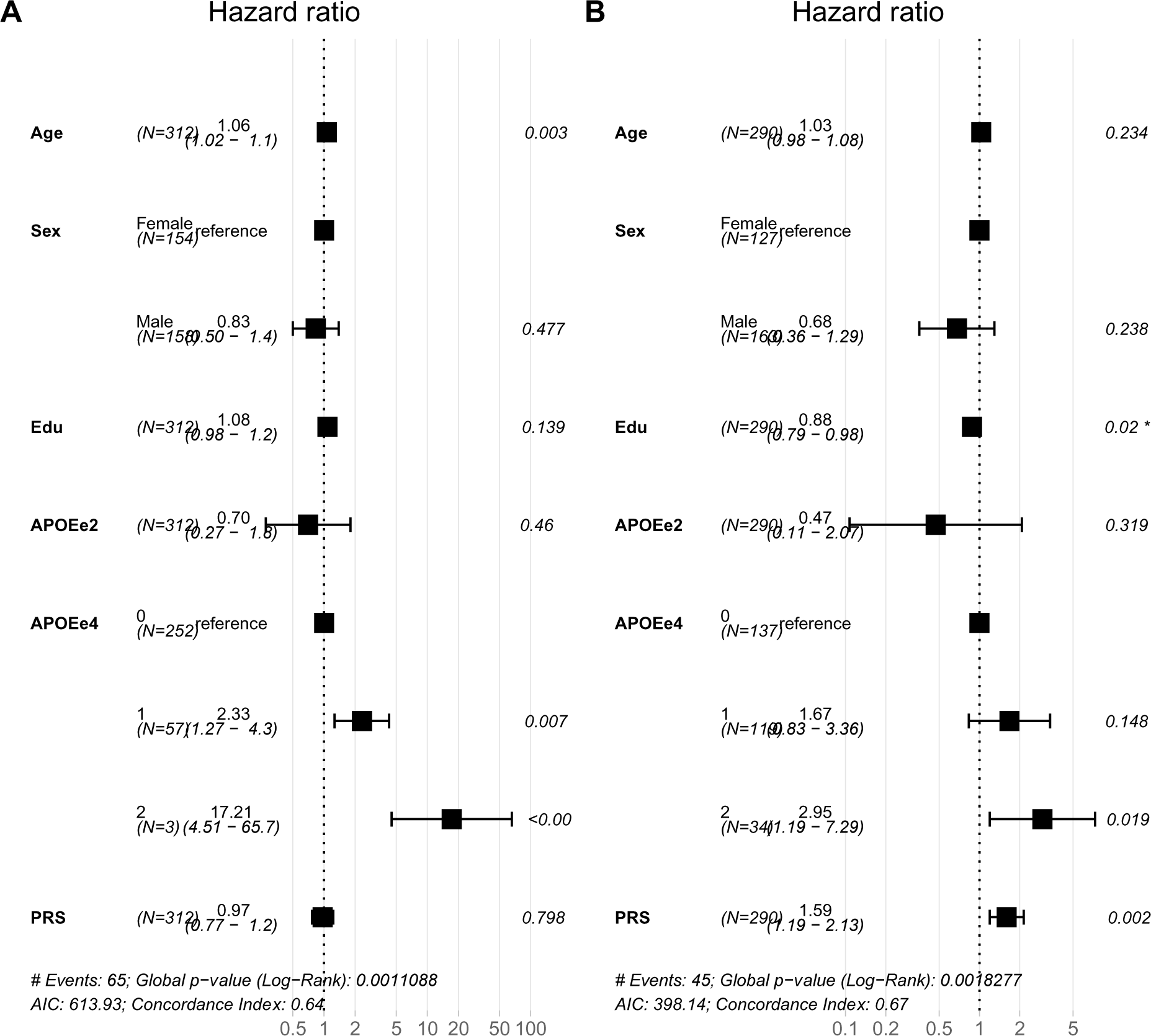
Hazard ratios for the PRS with 77 SNPs based on Bellenguez *et al.* (2022). (A) Forest plot depicting the Hazards Ratios (HR) for all covariates in the model for the A-T- to A+T- conversion. (B) Forest plot depicting the Hazards Ratios (HR) for all covariates in the model for the A+T- to A+T+ conversion. Edu = Years of Educations; APOEe2 = number of APOE2 alleles, APOEe4 = number of APOE4 alleles; PRS = polygenic risk score, scaled to zero mean and unit standard deviation.

**Supplementary Table 1:** Genetic variants contributing to the Polygenic Risk Score.

| Chr | Position | SNP | A1 | A2 | Gene | MAF | Weight | P-value |
| --- | --- | --- | --- | --- | --- | --- | --- | --- |
| 1 | 207750568 | rs679515 | T | C | *CR1* | 0.1778 | 0.1508 | 1.56E-16 |
| 2 | 127889932 | rs6710467 | A | G | *BIN1* | 0.1706 | 0.133 | 8.96E-12 |
| 2 | 127892810 | rs6733839 | T | C | *BIN1* | 0.3879 | 0.1693 | 4.02E-28 |
| 8 | 27219987 | rs73223431 | T | C | *PTK2B* | 0.3578 | 0.0936 | 8.34E-10 |
| 8 | 27468503 | rs867230 | C | A | *CLU* | 0.3958 | -0.1333 | 3.49E-17 |
| 11 | 47380340 | rs3740688 | G | T | *SPI1* | 0.4331 | -0.0935 | 9.70E-11 |
| 11 | 47663049 | rs10838738 | G | A | *MTCH2* | 0.3348 | -0.0871 | 5.99E-09 |
| 11 | 47915299 | rs34467936 | G | A | *PTPRJ* | 0.3372 | -0.0905 | 7.89E-09 |
| 11 | 60021948 | rs1582763 | A | G | *MS4A4A* | 0.3516 | -0.1232 | 1.19E-16 |
| 11 | 85868640 | rs3851179 | T | C | *PICALM* | 0.3443 | -0.1198 | 5.81E-16 |
| 14 | 92938855 | rs12590654 | A | G | *SLC24A4* | 0.3402 | -0.0906 | 8.73E-09 |
| 19 | 1047078 | rs4147910 | G | A | *ABCA7* | 0.1077 | 0.1488 | 9.27E-09 |
| 19 | 1050874 | rs12151021 | A | G | *ABCA7* | 0.3371 | 0.1071 | 2.56E-10 |

Chr: Chromosome; Position: position of the SNP in hg19; A1: effect allele; A2: reference allele; Gene: closest gene; MAF: minor allele frequency in the joint ADNI data ( $n=2001$ ); Weight: beta coefficient from the genome-wide association study (GWAS); P-value: p-value from the stage 1 GWAS by Kunkle et al. (2019).

**Supplementary Table 2:** Genetic variants contributing to the alternative Polygenic Risk Score.

| Chr | Position | SNP | A1 | A2 | Gene | EAF | Weight | P-value |
| --- | --- | --- | --- | --- | --- | --- | --- | --- |
| 1 | 161185602 | rs4575098 | A | G | ADAMTS4 | 0.2404 | 0.0539 | 1.33E-08 |
| 1 | 161217875 | rs2070902 | T | C | FCER1G | 0.2539 | -0.0537 | 1.05E-08 |
| 1 | 207221638 | rs6540874 | T | C | C4BPA | 0.7938 | -0.0706 | 1.70E-12 |
| 1 | 207518704 | rs6656401 | A | G | CR1 | 0.1881 | 0.1253 | 2.84E-33 |
| 2 | 127042739 | rs56049505 | T | G | BIN1 | 0.7987 | -0.0618 | 1.14E-09 |
| 2 | 127072128 | rs75836995 | T | C | BIN1 | 0.7892 | 0.0816 | 6.92E-16 |
| 2 | 127072643 | rs17014873 | A | C | BIN1 | 0.9293 | -0.1166 | 2.36E-13 |
| 2 | 127130409 | rs12989701 | A | C | BIN1 | 0.1522 | 0.0802 | 8.02E-13 |
| 2 | 127133508 | rs34546266 | A | G | BIN1 | 0.848 | -0.0762 | 2.64E-11 |
| 2 | 127135234 | rs6733839 | T | C | BIN1 | 0.3891 | 0.1686 | 6.48E-90 |
| 2 | 233117495 | rs7421448 | T | C | INPP5D | 0.0968 | -0.1091 | 1.01E-13 |
| 2 | 233160866 | rs28459768 | A | C | INPP5D | 0.5186 | -0.0608 | 1.31E-13 |
| 4 | 993555 | rs3822030 | T | G | IDUA | 0.5712 | 0.0514 | 5.04E-10 |
| 4 | 11023507 | rs6846529 | T | C | - | 0.7174 | -0.0673 | 1.25E-13 |
| 5 | 87002714 | rs62375397 | T | C | - | 0.2104 | 0.0746 | 8.58E-14 |
| 5 | 87111682 | rs2624182 | A | G | - | 0.7405 | 0.0517 | 3.94E-08 |
| 5 | 180201150 | rs113706587 | A | G | RASGEF1C | 0.1103 | 0.0925 | 3.38E-12 |
| 6 | 32373621 | rs9391858 | A | G | TSBP1 | 0.8475 | -0.068 | 2.79E-09 |
| 6 | 32408571 | rs3763312 | A | G | BTNL2 | 0.2036 | -0.0708 | 5.88E-12 |
| 6 | 32713500 | rs2858331 | A | G | HLA-DQA2 | 0.5891 | -0.0544 | 8.76E-11 |
| 6 | 32714360 | rs3957148 | A | G | HLA-DQA2 | 0.8996 | 0.1066 | 7.93E-15 |
| 6 | 41192063 | rs9394766 | A | G | TREML2 | 0.7013 | 0.0616 | 3.27E-12 |
| 6 | 41272084 | rs4714447 | T | C | TREM1 | 0.3449 | 0.0578 | 1.73E-11 |
| 6 | 47543755 | rs9349413 | A | G | CD2AP | 0.735 | -0.0649 | 1.61E-12 |
| 6 | 114361563 | rs976271 | A | G | HS3ST5 | 0.3613 | 0.0467 | 3.24E-08 |
| 7 | 28135367 | rs67250450 | T | C | JAZF1 | 0.7875 | 0.0559 | 2.02E-08 |
| 7 | 54881563 | rs74504435 | A | G | - | 0.9065 | 0.0842 | 2.05E-09 |
| 7 | 100045122 | rs4424195 | A | G | ZKSCAN1 | 0.7209 | 0.057 | 3.21E-10 |
| 7 | 100176704 | rs12705074 | A | G | GPC2 | 0.1121 | 0.0731 | 1.80E-08 |
| 7 | 100334426 | rs7384878 | T | C | PMS2P1 | 0.69 | 0.0775 | 2.13E-18 |
| 7 | 100489836 | rs13235951 | T | C | NYAP1 | 0.1266 | 0.0751 | 9.56E-10 |
| 7 | 100592493 | rs2734895 | T | C | FBXO24 | 0.3128 | -0.0586 | 6.58E-11 |
| 7 | 143413669 | rs11771145 | A | G | EPHA1 | 0.3476 | -0.0604 | 1.29E-12 |
| 8 | 27362470 | rs73223431 | T | C | PTK2B | 0.3694 | 0.0656 | 5.34E-15 |
| 8 | 27545260 | rs7341557 | A | G | EPHX2 | 0.1011 | -0.0986 | 8.78E-13 |
| 8 | 27562086 | rs11780834 | A | G | GULOP | 0.8236 | -0.0626 | 3.42E-09 |
| 8 | 27610986 | rs867230 | A | C | CLU | 0.6031 | 0.1009 | 1.50E-33 |
| 8 | 94988691 | rs4734295 | A | G | NDUFAF6 | 0.5394 | -0.0487 | 1.98E-09 |
| 8 | 100663356 | rs1693551 | T | C | SNX31 | 0.5328 | -0.0459 | 1.79E-08 |
| 10 | 11450869 | rs11257101 | T | C | USP6NL | 0.6898 | 0.0481 | 4.69E-08 |
| 10 | 11676714 | rs7912495 | A | G | ECHDC3 | 0.5381 | -0.0572 | 2.87E-12 |
| 10 | 60025170 | rs7068231 | T | G | ANK3 | 0.4026 | -0.0487 | 6.79E-09 |
| 10 | 80494228 | rs6586028 | T | C | TSPAN14 | 0.8036 | 0.0791 | 1.33E-14 |
| 11 | 47389337 | rs10838702 | T | G | SLC39A13 | 0.4008 | 0.0565 | 7.84E-12 |
| 11 | 47759202 | rs7927445 | T | G | FNBP4 | 0.3105 | 0.0498 | 1.29E-08 |
| 11 | 60254475 | rs1582763 | A | G | MS4A4A | 0.371 | -0.086 | 1.65E-24 |
| 11 | 86095220 | rs568755 | A | G | PICALM | 0.6909 | -0.05 | 1.23E-08 |
| 11 | 86152038 | rs56157503 | A | C | PICALM | 0.8768 | -0.0882 | 7.69E-13 |
| 11 | 86156833 | rs10792832 | A | G | PICALM | 0.3578 | -0.1056 | 6.33E-36 |
| 11 | 121487553 | rs60228070 | T | C | SORL1 | 0.8997 | -0.0756 | 2.70E-08 |
| 11 | 121566862 | rs1784920 | A | G | *SORL1* | 0.1215 | -0.0827 | 1.35E-10 |
| 14 | 39418472 | rs74745468 | A | G | *FBXO33* | 0.0875 | 0.0822 | 1.40E-08 |
| 14 | 52924962 | rs17125924 | A | G | *FERMT2* | 0.9108 | -0.0881 | 5.82E-10 |
| 14 | 92472511 | rs12590654 | A | G | *SLC24A4* | 0.3279 | -0.0693 | 2.08E-15 |
| 15 | 49901356 | rs2009833 | A | G | *ATP8B4* | 0.3683 | -0.0478 | 3.11E-08 |
| 15 | 58753575 | rs593742 | A | G | *ADAM10* | 0.7052 | 0.061 | 1.04E-11 |
| 15 | 63312881 | rs16946801 | T | C | *APH1B* | 0.6204 | 0.0622 | 1.15E-13 |
| 16 | 30022312 | rs12325539 | T | C | *DOC2A* | 0.6189 | 0.0567 | 1.25E-11 |
| 16 | 31143037 | rs78924645 | A | G | *PRSS36* | 0.2795 | -0.0565 | 7.71E-10 |
| 16 | 81739604 | rs12444183 | A | G | *PLCG2* | 0.3872 | -0.0591 | 2.21E-12 |
| 16 | 81942226 | rs4485362 | T | G | *PLCG2* | 0.5396 | 0.0463 | 2.00E-08 |
| 16 | 90103687 | rs56407236 | A | G | *FAM157C* | 0.0693 | 0.1097 | 1.28E-11 |
| 17 | 5220566 | rs113762960 | A | G | *SCIMP* | 0.129 | 0.0864 | 7.78E-13 |
| 17 | 44352876 | rs5848 | T | C | *GRN* | 0.2886 | 0.0646 | 1.76E-12 |
| 17 | 45846317 | rs12185268 | A | G | *SPPL2C<sup>a</sup>* | 0.774 | 0.0536 | 3.85E-08 |
| 17 | 46107462 | rs4510068 | T | G | *KANSL1<sup>a</sup>* | 0.3938 | -0.0546 | 1.37E-10 |
| 17 | 46720553 | rs199456 | T | C | *NSF<sup>a</sup>* | 0.2123 | -0.0593 | 2.57E-09 |
| 17 | 58363181 | rs2680700 | T | G | *RNF43* | 0.3755 | -0.0482 | 9.90E-09 |
| 17 | 63476980 | rs4292 | T | C | *ACE* | 0.617 | 0.0688 | 3.47E-16 |
| 19 | 1032690 | rs1141534 | T | G | *CNN2* | 0.204 | 0.073 | 3.65E-11 |
| 19 | 1047079 | rs4147910 | A | G | *ABCA7* | 0.8995 | -0.0977 | 1.47E-11 |
| 19 | 1050875 | rs12151021 | A | G | *ABCA7* | 0.3357 | 0.1055 | 4.09E-30 |
| 19 | 1085456 | rs72975514 | T | C | *ARHGAP45* | 0.7924 | 0.06 | 4.60E-08 |
| 19 | 1833339 | rs732310 | T | G | *REXO1* | 0.5158 | -0.05 | 1.33E-08 |
| 20 | 56423488 | rs6014724 | A | G | *CASS4* | 0.9102 | 0.1176 | 4.84E-16 |
| 21 | 26101558 | rs2154481 | T | C | *APP* | 0.5236 | 0.05 | 1.02E-09 |
| 21 | 26775872 | rs2830489 | T | C | *ADAMTS1* | 0.2809 | -0.0547 | 1.72E-09 |

Chr: Chromosome; Position: position of the SNP in hg38; A1: effect allele; A2: reference allele; Gene: closest protein-coding gene within 100kb; EAF: effect allele frequency; Weight: beta coefficient from the genome-wide association study (GWAS); P-value: p-value from the stage I GWAS by Bellenguez *et al.* (2022). <sup>a</sup>: *MAPT* locus

**Supplementary Table 3:** Demographics (relaxed conversion criterion).

|  | **A-T-** |  |  | **A+T-** |  |  |
| --- | --- | --- | --- | --- | --- | --- |
|  | **Total** | **Stable** | **Converter** | **Total** | **Stable** | **Converter** |
| N | 400 | 315 | 85 | 311 | 260 | 51 |
| Age, y (sd) | 71.0 (6.9) | 70.8 (6.7) | 72.1 (7.1) | 73.2 (7.0) | 73.1 (7.0) | 73.5 (7.0) |
| Sex (%female) | 48.0 | 47.6 | 49.4 | 43.1 | 42.7 | 45.1 |
| DX (CN/MCI/AD) | 229/166/5 | 174/137/4 | 55/29/1 | 131/161/19 | 112/131/17 | 19/30/2 |
| Education, y (sd) | 16.7 (2.4) | 16.6 (2.5) | 17.2 (2.2) | 16.5 (2.6) | 16.6 (2.6) | 15.8 (2.4) |
| Years Follow-up, y (sd) | 4.9 (3.0) | 4.6 (2.8) | 6.3 (3.4) | 4.0 (2.7) | 3.7 (2.6) | 5.5 (2.4) |
| Time to event, y (sd) | n/a | n/a | 4.2 (2.7) | n/a | n/a | 4.0 (2.4) |
| APOE-e4 (0/1/2) | 321/76/3 | 266/49/0 | 55/27/3 | 148/126/37 | 129/103/28 | 19/23/9 |
| APOE-e2 (0/1/2) | 347/52/1 | 268/46/1 | 79/6/0 | 291/20/0 | 242/18/0 | 49/2/0 |
| PRS (sd) | 0.013 (0.012) | 0.013 (0.012) | 0.013 (0.011) | 0.014 (0.013) | 0.013 (0.013) | 0.019 (0.012) |

DX = diagnosis (cognitively normal [CN]; mild cognitive impairment [MCI]; Alzheimer's Disease (AD)); PRS = Polygenic Risk Score

**Supplementary Table 4:** Sensitivity analysis for different Tau PET cutoffs.

|  | Tau PET | CSF pTau | n | Events | APOE e4 | APOE e4/4 | PRS |
| --- | --- | --- | --- | --- | --- | --- | --- |
| A-T- to A+T- | 1.31 | 32.30 | 309 | 62 | 0.02915643 | 4.06E-05 | 0.6499623 |
|  | 1.34 | 32.95 | 310 | 64 | 0.01775002 | 3.55E-05 | 0.5021401 |
|  | 1.37 | 33.57 | 311 | 65 | 0.00726623 | 2.20E-05 | 0.4358987 |
|  | 1.39 | 34.13 | 311 | 65 | 0.00699722 | 2.02E-05 | 0.5160777 |
|  | **1.42** | **34.61** | **312** | **65** | **0.00639089** | **1.81E-05** | **0.4962599** |
|  | 1.45 | 34.82 | 314 | 67 | 0.00904438 | 1.41E-05 | 0.5737404 |
|  | 1.48 | 35.07 | 315 | 67 | 0.00896343 | 1.39E-05 | 0.5734747 |
|  | 1.51 | 35.29 | 315 | 67 | 0.00896343 | 1.39E-05 | 0.5734747 |
|  | 1.54 | 35.53 | 316 | 67 | 0.00885485 | 1.38E-05 | 0.5735298 |
| A+T- to A+T+ | 1.31 | 32.30 | 258 | 46 | 0.05796633 | 0.48549513 | 0.00204043 |
|  | 1.34 | 32.95 | 266 | 44 | 0.10834803 | 0.2154368 | 0.00084655 |
|  | 1.37 | 33.57 | 271 | 42 | 0.06722642 | 0.04442165 | 0.00097163 |
|  | 1.39 | 34.13 | 280 | 43 | 0.07594692 | 0.09580913 | 0.00069806 |
|  | **1.42** | **34.61** | **290** | **45** | **0.11930622** | **0.03939911** | **0.0005569** |
|  | 1.45 | 34.82 | 290 | 43 | 0.18026366 | 0.02915812 | 0.00089779 |
|  | 1.48 | 35.07 | 291 | 39 | 0.55440604 | 0.02044481 | 0.0076896 |
|  | 1.51 | 35.29 | 293 | 41 | 0.30800169 | 0.02089209 | 0.0048998 |
|  | 1.54 | 35.53 | 296 | 39 | 0.37466685 | 0.02998641 | 0.00162742 |

Tau PET: Tau PET threshold based on 1.0 to 3.0 standard deviations (in steps of 0.25) above the mean in cognitively normal participants. CSF pTau: pTau threshold that maximized the Youden's index for the corresponding Tau PET cutoff. n: sample size; Events: number of converters; APOE e4: p-value for APOEe4 heterozygotes; APOE e4/4: p-value for APOEe4 homozygotes; PRS: p-value for the Polygenic Risk Score. Bold font indicates the settings used for the main analysis (i.e., Tau PET cutoff at a z-score of 2.0).

**Supplementary Table 5:** Sensitivity analysis using only Tau PET.

|  | Tau PET | *n* | Events | APOE e4 | APOE e4/4 | PRS |
| --- | --- | --- | --- | --- | --- | --- |
| A-T- to A+T- | 1.31 | 108 | 15 | 0.00256332 | 0.998414 | 0.5466185 |
|  | 1.34 | 110 | 17 | 0.00617315 | 0.998169 | 0.375093 |
|  | 1.37 | 112 | 18 | 0.00194 | 0.9983591 | 0.3558922 |
|  | 1.39 | 113 | 18 | 0.0019078 | 0.9983644 | 0.3514293 |
|  | 1.42 | 113 | 18 | 0.0019078 | 0.9983644 | 0.3514293 |
|  | **1.45** | **113** | **19** | **0.00276536** | **0.9982344** | **0.5255481** |
|  | 1.48 | 113 | 19 | 0.00276536 | 0.9982344 | 0.5255481 |
|  | 1.51 | 113 | 19 | 0.00276536 | 0.9982344 | 0.5255481 |
|  | 1.54 | 113 | 19 | 0.00276536 | 0.9982344 | 0.5255481 |
| A+T- to A+T+ | 1.31 | 70 | 9 | 0.4401025 | 0.9988313 | 0.1140501 |
|  | 1.34 | 73 | 8 | 0.3714046 | 0.9987921 | 0.21299683 |
|  | 1.37 | 78 | 9 | 0.2152745 | 0.2792531 | 0.15481342 |
|  | 1.39 | 80 | 7 | 0.5417669 | 0.2673798 | 0.02184563 |
|  | 1.42 | 90 | 13 | 0.4710315 | 0.6737454 | 0.01884522 |
|  | **1.45** | **91** | **13** | **0.6017861** | **0.8375521** | **0.01031391** |
|  | 1.48 | 91 | 10 | 0.2393688 | 0.4005217 | 0.04235292 |
|  | 1.51 | 92 | 11 | 0.3619229 | 0.3545669 | 0.03555231 |
|  | 1.54 | 94 | 12 | 0.14025 | 0.1759473 | 0.00334521 |

Tau PET: Tau PET threshold based on 1.0 to 3.0 standard deviations (in steps of 0.25) above the mean in cognitively normal participants. *n*: sample size; Events: number of converters; APOE e4: p-value for APOEe4 heterozygotes; APOE e4/4: p-value for APOEe4 homozygotes; PRS: p-value for the Polygenic Risk Score. Bold font indicates the settings presented in the main analysis (i.e., Tau PET cutoff at a z-score of 1.0).

**Supplementary Table 6:**
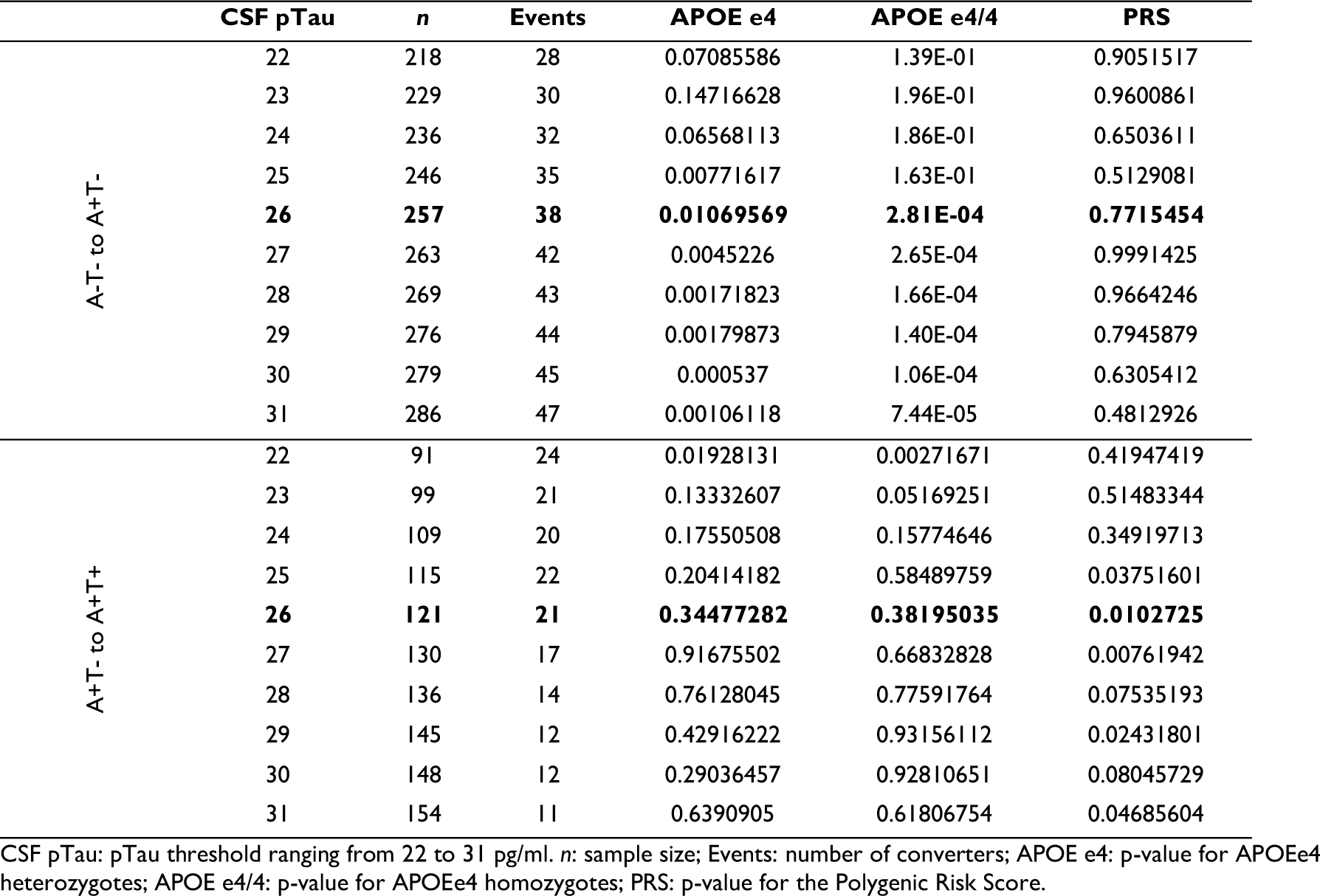
Sensitivity analysis using only CSF pTau.

|  | CSF pTau | n | Events | APOE e4 | APOE e4/4 | PRS |
| --- | --- | --- | --- | --- | --- | --- |
| A-T- to A+T- | 22 | 218 | 28 | 0.07085586 | 1.39E-01 | 0.9051517 |
|  | 23 | 229 | 30 | 0.14716628 | 1.96E-01 | 0.9600861 |
|  | 24 | 236 | 32 | 0.06568113 | 1.86E-01 | 0.6503611 |
|  | 25 | 246 | 35 | 0.00771617 | 1.63E-01 | 0.5129081 |
|  | **26** | **257** | **38** | **0.01069569** | **2.81E-04** | **0.7715454** |
|  | 27 | 263 | 42 | 0.0045226 | 2.65E-04 | 0.9991425 |
|  | 28 | 269 | 43 | 0.00171823 | 1.66E-04 | 0.9664246 |
|  | 29 | 276 | 44 | 0.00179873 | 1.40E-04 | 0.7945879 |
|  | 30 | 279 | 45 | 0.000537 | 1.06E-04 | 0.6305412 |
|  | 31 | 286 | 47 | 0.00106118 | 7.44E-05 | 0.4812926 |
| A+T- to A+T+ | 22 | 91 | 24 | 0.01928131 | 0.00271671 | 0.41947419 |
|  | 23 | 99 | 21 | 0.13332607 | 0.05169251 | 0.51483344 |
|  | 24 | 109 | 20 | 0.17550508 | 0.15774646 | 0.34919713 |
|  | 25 | 115 | 22 | 0.20414182 | 0.58489759 | 0.03751601 |
|  | **26** | **121** | **21** | **0.34477282** | **0.38195035** | **0.0102725** |
|  | 27 | 130 | 17 | 0.91675502 | 0.66832828 | 0.00761942 |
|  | 28 | 136 | 14 | 0.76128045 | 0.77591764 | 0.07535193 |
|  | 29 | 145 | 12 | 0.42916222 | 0.93156112 | 0.02431801 |
|  | 30 | 148 | 12 | 0.29036457 | 0.92810651 | 0.08045729 |
|  | 31 | 154 | 11 | 0.6390905 | 0.61806754 | 0.04685604 |

CSF pTau: pTau threshold ranging from 22 to 31 pg/ml. n: sample size; Events: number of converters; APOE e4: p-value for APOEe4 heterozygotes; APOE e4/4: p-value for APOEe4 homozygotes; PRS: p-value for the Polygenic Risk Score.

